# An Evaluation of Liver Function Tests in SARS-CoV-2 infection in the backdrop of chronic kidney disease

**DOI:** 10.1101/2021.07.16.21260406

**Authors:** Rajeev Kumar, Pratip Jana, Indu Priyadarshini, Smita Roy, Pritha Datta, Saswati Das

## Abstract

**INTRODUCTION:** The SARS-CoV-2 pandemic has emerged as perhaps the most challenging global health problem of this century. The concomitant presence of co-morbidities like chronic kidney disease (CKD), diabetes, chronic heart disease etc. makes the task of patient management difficult.

**AIMS AND OBJECTIVES:** To assess the patterns of liver test abnormalities in patients of COVID-19 infection with and without CKD and evaluate the probable outcomes.

**MATERIALs and METHODS:** A cross-sectional retrospective observational study done on 600 patient samples (Group 1 COVID-19 without CKD, Group 2 COVID-19 with CKD and Group 3 non COVID-19 with CKD) which were processed for Liver function test (AST, ALT and ALP) and Renal function test (Urea and Creatinine) in the Department of Biochemistry, Dr. RML Hospital New Delhi.

**RESULTS:** AST and ALT levels were significantly higher (P < 0.05) in all COVID-19 positive patients - group 1 mean ± 2 SD, (63.63 ± 42.89 U/L & 50.25 ± 46.53 U/L respectively) and group 2 (90.59 ± 62.51 U/L & 72.09 ± 67.24 U/L respectively) as compared to Group 3 (25.24 ± 7.47 U/L & 24.93 ± 11.44 U/L respectively) and also a statistically significant elevation is seen in these two parameters (AST & ALT) in Group 2 as compared to Group 1 (*P* < 0.05). There was a negative significant correlation between eGFR and AST/ALT levels in Group 1 (p < 0.05). In Group 2, a weak positive correlation was seen with ALT (p < 0.01).

No significant correlation existed between eGFR and ALP in groups 1 and 2. In Group 3, eGFR’s showed strong correlations with AST and ALT levels (p < 0.01) and reduction in kidney function correlated well with increase in serum ALP levels, (p < 0.01).

**CONCLUSIONS:** This study most comprehensively describes that SARS-CoV-2 positive CKD patients show more elevations in serum aminotransferase levels as compared to their non-CKD counterparts, in contrast to non-COVID-19 CKD cases. Serum ALT values in SARS-CoV-2 patients show significant correlation with calculated eGFR values. Elevated ALP values in CKD patients may be used as an indicator of declining kidney function. However, more studies in this direction are needed.

## INTRODUCTION

The SARS-CoV-2 pandemic commonly referred as COVID-19 disease has emerged as the most challenging global health problem of this century. Since its first emergence as a pneumonia of unknown cause, in Wuhan City, on the last day of 2019 and subsequent identification as the cause by Chinese authorities on 7^th^ January, 2020^1^ it has ravaged all continents and left a huge trail of death and devastation in its path. Initially christened as the novel coronavirus, the International Committee on Taxonomy of Viruses adopted the official name “severe acute respiratory syndrome coronavirus 2” (SARS-CoV-2) on 11 February 2020.^2^ World Health Organization figures say, it has infected more than 100 million people and claimed more than 2.15 million lives worldwide. As on 07 June 2021, India reported 3.06 Cr confirmed cases and total of 4.03 L deaths have been reported.^3^

SARS-CoV-2 infection is primarily a disease of respiratory system. Although the pulmonary manifestations are the most common presentation, important extra-pulmonary events include gastrointestinal complications, cardiovascular injury, and renal dysfunction^4-7^.

The concomitant presence of co-morbidities like chronic kidney disease (CKD)-encompasses a huge gamut of conditions associated with a progressive decline in kidney functions and abnormal glomerular filtration rate, makes the task of patient management a daunting challenge for the attending physician.

The objective of this study is to compare the levels of the liver enzymes serum aspartate aminotransferase (AST), alanine aminotransferase (ALT) and serum alkaline phosphatase (ALP) levels among the three groups of subjects. Group 1 comprising of COVID-19 RT-PCR positive patients without CKD, Group 2 COVID-19 RT-PCR positive patients with CKD, and Group 3 COVID-19 RT-PCR negative CKD patients. In our paper we systematically assess the patterns of liver test abnormalities in patients of COVID-19 infection with and without CKD, and further evaluate the probable outcome on the basis of these findings.

### Methodology

This is a cross-sectional retrospective observational study conducted at the Department of Biochemistry, Atal Bihari Vajpayee Institute of Medical Sciences & Dr. Ram Manohar Lohia Hospital, New Delhi, India. The data was collected from laboratory reports obtained using the hospital database, by biochemical analysis of blood samples, from patients admitted to the different COVID-19 designated wards/ICUs and Nephrology Unit of Atal Bihari Vajpayee Institute of Medical Sciences & Dr. Ram Manohar Lohia Hospital, New Delhi, a tertiary care teaching center. Data from the month of July 2020 to November 2020 was analyzed.

A total of 600 patient samples were included in the study.454 RT-PCR positive SARS-Cov-2 subjects, of age ≥18 years and of either sex, were included in the study. Among the study subjects 253 were cases of COVID-19 without altered renal function and 201 of COVID-19 with pre-existing renal disease (CKD stage 3 and 4). A second cohort of 219 non-COVID-19 cases (negative RT-PCR for SARS-Cov-2) with Stage 3 & 4 CKD of similar demographic profile, were included in this study. The first 200 consecutive patient records in ascending order of their patient unique identification number in each group were included in the study. Cases with active COVID-19 infection from COVID-19 designated ward/ICUs were divided into two groups as those without CKD (Group 1) and with CKD (Group 2). Non COVID-19 cases with CKD from Nephrology OPD have been included in Group 3.

### Inclusion and exclusion criteria are listed as below

**Inclusion and exclusion criteria are listed as below:**
| INCLUSION CRITERIA | GROUPS |  |  |
| --- | --- | --- | --- |
|  | GROUP 1 | GROUP 2 | GROUP 3 |
| **1. Patients who were symptomatic and tested positive for SARS-CoV-2 by RT-PCR test** | + | + | - |
| **2. Sex. (Male/Female)** | M/F | M/F | M/F |
| **3. Age $\geq 18$ years** | + | + | + |
| **4.History of chronic kidney disease (CKD)** | – | + | + |

Exclusion criteria - Pregnant and lactating females, patients on hemo-dialysis/ESRD as well as patients with previous history of CLD or preexisting cardiac disease (CHD)

All samples were collected adhering to bio-safety guidelines and received according to ICMR 2019 guidelines of COVID-19 management, in plain vacutainers with triple layer packaging. They were processed for liver function tests (LFT) - AST [ULN (upper limit of normal)-41U/L], ALT [ULN-40U/L], ALP [ULN-135 U/L] and renal function tests (Serum Urea and Creatinine) in COVID-19 (Trauma center) Laboratory, Department of Biochemistry, Dr. RML Hospital New Delhi. The cases with elevated liver enzyme values were considered to have abnormal liver function. Those with ALT, AST values > 3xULN, and ALP>2.5xULN, were classified as those having liver injury. Serum AST & ALT were estimated by ERBA Mannheim XL system Pack IFCC recommended without pyridoxal phosphate reagent; serum ALP was estimated by using ERBA Mannheim XL system Pack German society of clinical chemistry recommended ALP AMP method; serum Urea was estimated by urease GLDH method and serum creatinine was estimated by enzymatic method on the venous blood samples after separation of serum by proper centrifugation. The samples were processed on fully automated ERBA XL 640 Chemistry Analyzer (TRANSASIA Bio-medicals, India) after verification of two levels of internal quality controls. The present study was approved by the Institutional Ethics Committee [416(65/2020)/IEC/ABVIMS/RMLH/222].

#### Statistical analysis

The data were analyzed using SPSS statistical software (v 24; IBM Corporation, Armonk, NY, USA). The data was checked for normality using Shapiro-Wilk’s test. Data is represented as mean and standard deviation for the serum levels of AST, ALT and ALP in all the three groups. The means of the different parameters in three groups were compared by one-way analysis of variance (ANOVA). These comparisons were followed by *post-hoc* comparisons between groups by means of the Tukey’s test and a *P* < 0.05 was considered statistically significant. The gender distribution was also compared among the three groups using Chi-square test. The linear relationship between the variables (test parameters) was assessed using the Spearman Rank Correlation analysis method. At 95% confidence interval, a p value of ≤ 0.05 was considered significant.

## RESULT

The three groups age and sex matched. There was no statistically significant difference (P > 0.05) between the mean ages in the three groups. No significant difference (P > 0.05) was observed between the percentages of females in the three groups as shown in Table no 1.

**Table 1:** Demographic characteristics of the three study Groups-1, 2, 3 (Group 1 COVID-19 without CKD, Group 2 COVID-19 with CKD, Group 3 Non-COVID-19 with CKD)

| Demographic parameters | Group 1 | Group 2 | Group 3 | p value |
| --- | --- | --- | --- | --- |
| Age in years<br>(mean ± SD) | 49.29 ± 18.52 | 54.84 ± 15.01 | 53.61 ± 14.49 | 0.724 |
| Range in years | 18 - 77 | 21 - 79 | 29–75 |  |
| Female (%) | 37.8 | 42.2 | 36.7 | 0.578 |

Of the 400 patients with COVID-19 whose test reports were analyzed, 61.5 (%) had abnormal liver function and 11.5 (%) had liver injury during hospitalization.

Table no 2 and 3 showing statistical comparisons of the means of the three groups made by one way ANOVA, followed by post-hoc analysis. A value of (P < 0.05) of the mean difference for the parameters in the 3 groups was considered to be significant.

**Table 2:** LFT outcomes amongst the 3 groups:

| Test Parameters (U/L) | GROUP 1<br>(n=200) | GROUP 2<br>(n=200) | GROUP 3<br>(n=200) |
| --- | --- | --- | --- |
| | Mean ± SD | Mean ± SD | Mean ± SD |
| AST | 63.63 ± 42.89 | 90.59 ± 62.51 | 25.24 ± 7.47 |
| ALT | 50.29 ± 46.53 | 72.09 ± 67.24 | 24.93 ± 11.44 |
| ALP | 123.39 ± 78.31 | 185.38 ± 92.70 | 186.22 ± 76.29 |

**Table 3:**
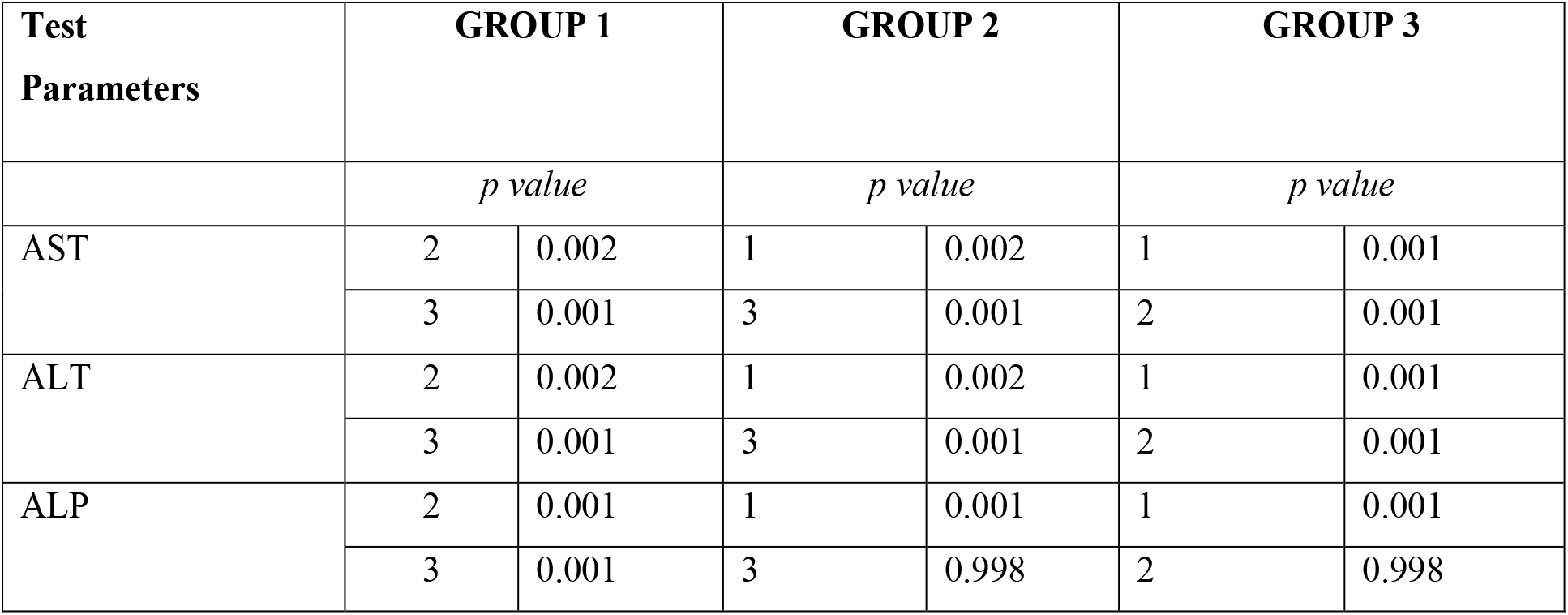
Correlation between LFT outcomes amongst the 3 groups:

As described in Table no 2 and 3 AST levels were significantly higher in all COVID-19 positive patients irrespective of their renal function status. AST levels were significantly higher in both Group 1 mean ± 2 SD (63.63 ± 42.89 U/L) and Group 2 (90.59 ± 62.51 U/L) as compared to Group 3 (25.24 ± 7.47 U/L) (P < 0.05). There was also a statistically significant elevation of serum AST levels in patients belonging to Group 2 as compared to patients in Group 1 (*P = 0*.*002*).

**Figure 1:**
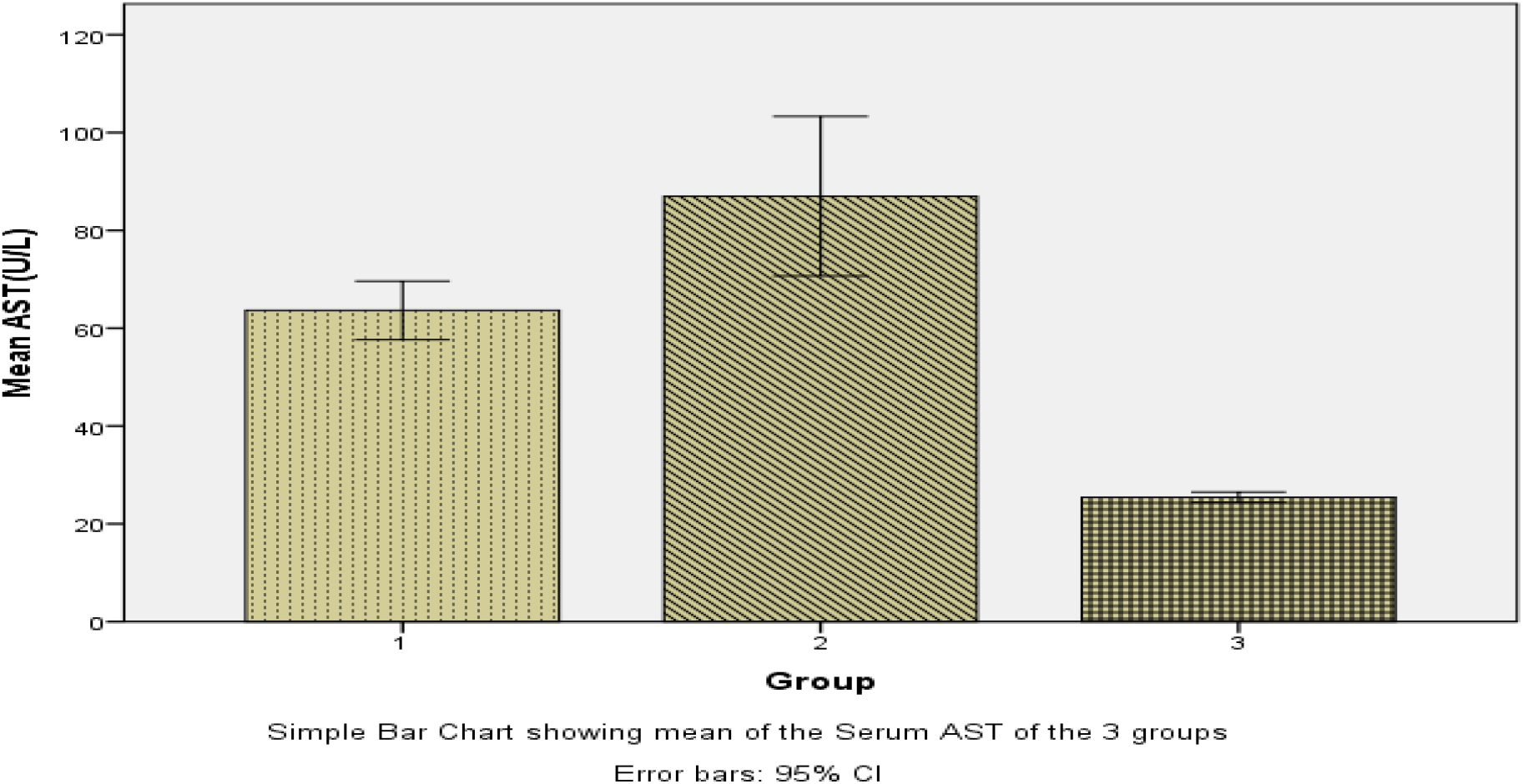
Simple Bar Chart showing mean of the Serum AST values in the 3 groups.

Similarly, serum ALT levels were also significantly higher in both Group 1, mean ± 2 SD (50.25 ± 46.53 U/L) and Group 2 (72.09 ± 67.24 U/L) as compared to Group 3 (24.93 ± 11.50 U/L) (P < 0.05). There was a significant difference seen between groups 1, group 2, and group 3 (p = 0.001, 0.002, 0.001) respectively.

**Figure 2:**
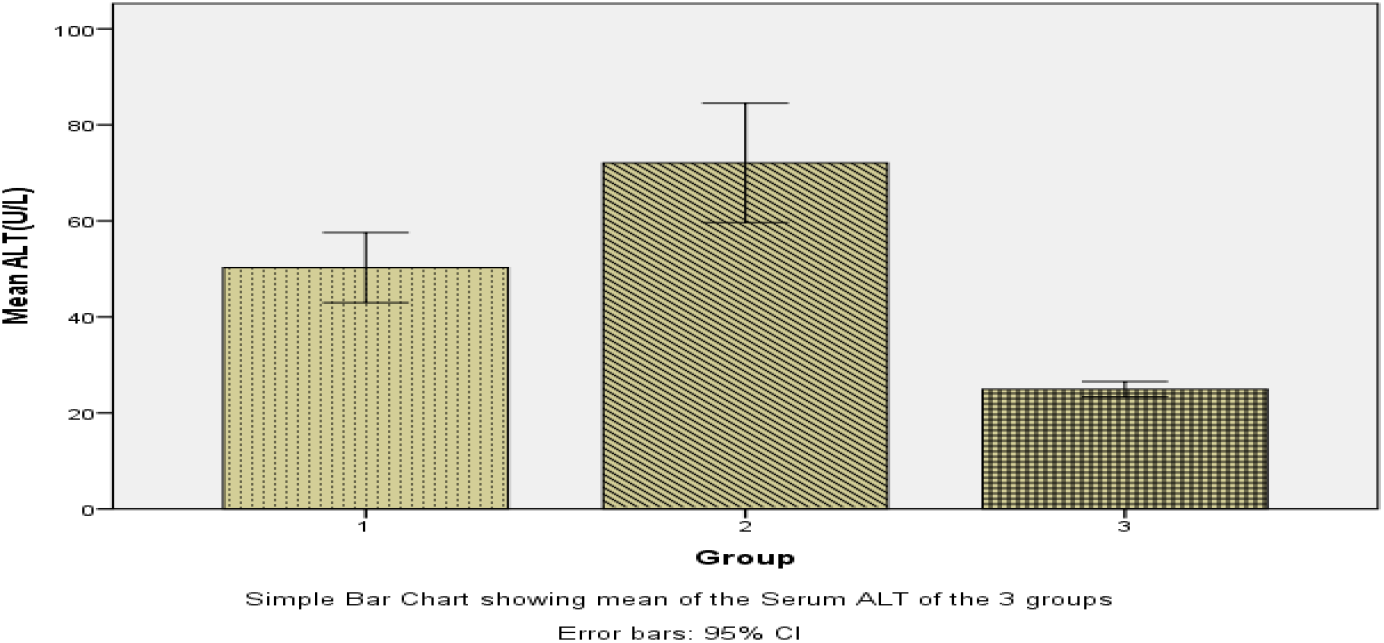
Simple Bar Chart showing mean of the Serum ALT values in the 3 groups.

On the contrary serum ALP levels were significantly lower in Group 1, mean ± 2 SD, (123.39 ± 78.31 U/L) (p < 0.05) as compared to Group 2 (185.38 ± 92.70 U/L) and Group 3 (186.22 ± 76.29 U/L). The average serum ALP levels of Groups 2 & 3 were more or less comparable, and no significant difference was seen (p = 0.998).

**Figure 3:**
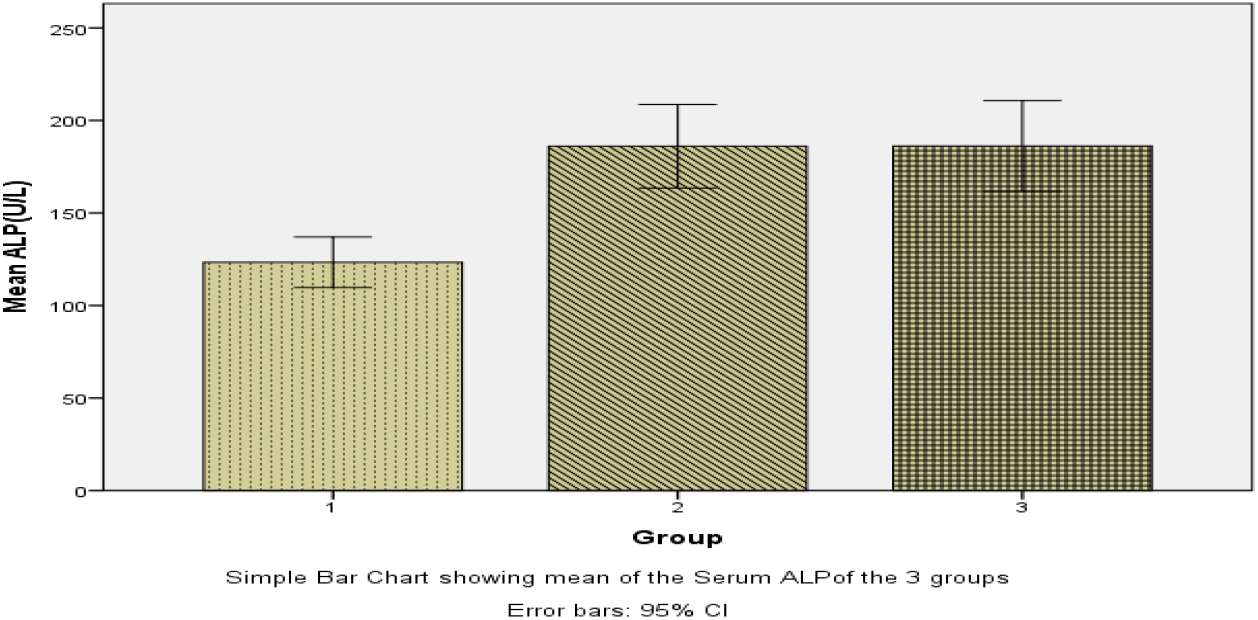
Simple Bar Chart showing mean of the Serum ALP values in the 3 groups.

Correlation studies between eGFR values and liver enzyme levels was performed in all the three groups as represented in table no 4. There was a negative correlation between eGFR and AST levels in Group 1 (Spearman’s rho =0.017 and p < 0.05). A similar but stronger trend was observed in case of ALT (rho = 0.001, p < 0.01). However no significant correlation existed between eGFR and ALP. In Group 2, no statistically significant correlation was seen between eGFR and AST or ALP values. However, a weak positive correlation was seen with ALT (rho =0.006, p < 0.01). In Group 3, eGFR showed strong positive correlations with AST and ALT levels (rho = 0.001, p < 0.01; rho= 0.001, p < 0.01) while reduction in kidney function correlated well with increase in serum ALP levels, (rho = 0.001, p < 0.01).

**Table 4:**
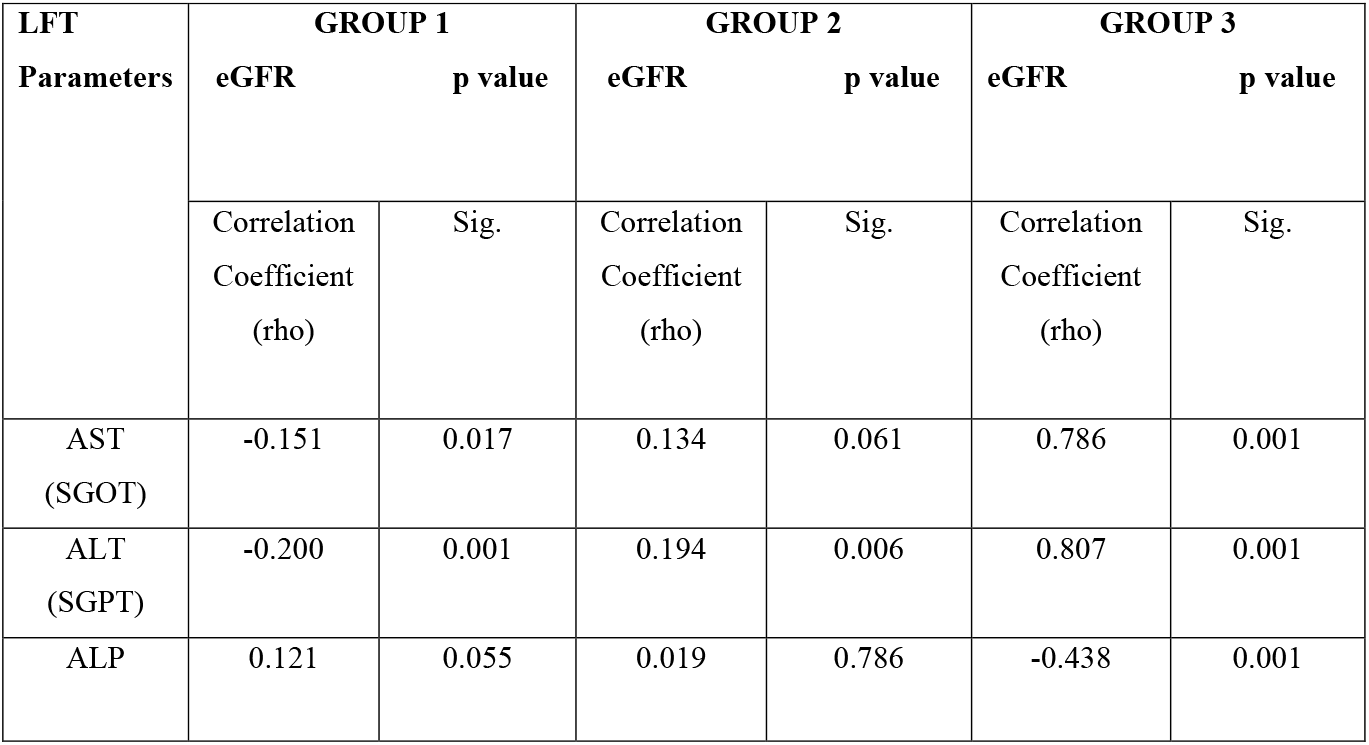
Correlation between eGFR and LFT amongst the 3 group.

## DISCUSSION

CKD is classified by the National Kidney Foundation guidelines, into 5 stages. This is based on the calculation of estimated GFR (eGFR)^8^. The Modification of Diet in Renal Disease (MDRD) Study equation is the most widely used equation for estimating GFR in patients aged 18 yrs. and over.^9^

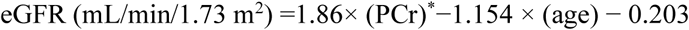

[Multiplied by 0.742 for women and by 1.21 for African Americans. PCr*-Plasma Creatinine.]

In Stage 1 eGFR is ≥ 90 mL/min/1.73 m^2^ with presence of kidney damage like persistent proteinuria, abnormal blood, and urine chemistry etc. Stages 2, 3 and 4 correspond to eGFR of 60 – 89 mL/min/1.73 m^2^, 30 – 59 mL/min/1.73 m^2^ and 15 – 29 mL/min/1.73 m^2^, respectively. The prime objective of treatment is to retard the progression of CKD, perform cardiovascular disease risk estimation and manage the complications^.9^ CKD stage 5 corresponds to eGFR 15mL/min/1.73 m^2^ and is also known as end stage renal disease (ESRD).

In our study, CKD stage 3 and 4 patients were considered in Groups 2 & 3. The association of co-morbidities is a characteristic of chronic kidney disease. Derangements of hepatic function is one of the most important co-morbid conditions commonly seen with CKD and may be particularly confounding in cases with associated COVID-19. Numerous studies have shown that hepatic enzymes levels act as good prognostic markers in CKD including ESRD.^10-11^ Several studies have concluded, that, there is a decrease in the level of serum amino-transferases in patients with CKD compared to the normal population. Ray L. et al concluded that serum aminotransferase levels tend to remain lower in ESRD patients compared to the normal population, and levels further reduce with the worsening of CKD.^10^

The pathophysiological mechanism for the reduction in the serum aminotransferase levels in patients with CKD remains controversial. The possible mechanisms include reduction in pyridoxal-5-phosphate which is a coenzyme of aminotransferase, presence of ultraviolet absorbing materials, and high levels of uremic toxins^10^. Other possibilities included decreased synthesis and inhibition of release of AST and ALT from hepatocytes or accelerated clearance from serum.^13-16^ A low serum aminotransferase level could also be due to water retention and hemodilution in patients of CKD.^10^ However, this pattern is usually not maintained in CKD with COVID-19. In our study we included patients from this group.

Different meta-analytical and independent studies have shown, that, more than half of patients with COVID-19 (SARS-CoV-2) showed varying levels of liver involvement. Several case reports have also indicated that a significant number amongst them show evidence of liver damage too.^17-19^About 30% of all COVID-19, RT PCR positive admissions in our hospital, had abnormally high levels of serum creatinine at the time of presentation. This intensive derangement of renal function may have been due to the acute deterioration of renal function related to COVID-19 (SARS-CoV-2) disease or increased susceptibility to secondary infection in them.^**20**^ Other possible reasons for the high prevalence of kidney involvement, is that some of the patients with COVID-19 infection, had a previously documented history of CKD.

Liver damage in patients with coronavirus infection might be directly caused by the viral infection of liver cells.^21^ Two recent studies showed that angiotensin converting enzyme 2 (ACE2) was the key receptor for COVID-19 (SARS-CoV-2) cell entry.^21-23^ which was mainly localized in the heart, kidney and testes, and expressed at a low level in many other tissues, especially in the colon and lung.^24^ A study showed that COVID-19 (SARS-CoV-2) might directly bind to ACE2 positive cholangiocytes and cause liver damage, which may partially explain the contribution of SARS-CoV-2 infection to the liver test dysfunction in our patients. The virus may bind to angiotensin converting enzyme 2 (ACE2) cholangiocytes, leading to cholangiocytes dysfunction and inducing a systemic inflammatory response leading to liver injury.^23,25^ Moreover, the use of ACE-inhibitors and angiotensin receptor blocker (ARBs) drugs might also affect liver tests.^26^ Another possible reason behind this, is the use of hepatotoxic drugs like the antiviral remdesivir, oxidative stress, coexisting systemic inflammatory response, respiratory distress induced hypoxia and associated multi-organ dysfunction.^17,27,28^

In our study serum AST levels were significantly higher in all COVID-19 positive patients irrespective of their renal function status. Also, there was a statistically significant elevation in Group 2 as compared to Group 1 (*P* < 0.05). Similarly, serum ALT levels were also significantly, higher in both Group 1, and Group 2 as compared to Group 3 (P < 0.05). Also, there was a significant difference seen between group 1 and group 2 (p< 0.05). Thus, the findings of serum AST and ALT levels are not in consonance with those seen in non-COVID-19 CKD patients and show an increase rather than a decrease. However, serum ALP levels were significantly lower in Group 1 as compared to Group 2 and 3, thereby indicating a higher strength of association of ALP values with CKD than non-CKD cases. The average serum ALP level of Group 2 and 3 were comparable. ALP levels in both the latter groups show a rising trend with a marginal increase in the mean of group 3 (185.38 ± 92.70 U/L) in comparison to group 2 (186.22 ± 76.29 U/L).

In Group 1 a negative correlation is seen between eGFR and AST (p < 0.05). A similar but stronger trend was observed in case of ALT (p < 0.01). In Group 2, both AST and ALT showed a weak positive correlation with eGFR; it was statistically significant only with the latter (ALT) (p < 0.01), signifying a reduction in ALT levels with deteriorating kidney function, in the COVID-19 positive CKD cases. A positive correlation existed between eGFR and ALP in groups 1 and 2, but this was not significant. In Group 3, eGFRs showed strong positive correlations with AST and ALT levels (p < 0.01) and reduction in kidney function correlated well with increase in serum ALP levels, (p < 0.01).

The association between ALP and renal damage may be, at least in part, explained by endothelial dysfunction, a strong and independent predictor of cardiovascular events in different clinical conditions, including essential hypertension^29^ which is one of the leading causes of CKD stage 4. Mechanisms linking ALP to endothelial dysfunction may include inhibition of tyrosine kinase activity into endothelial cells with consequent impairment of endothelial NO synthase function, promotion of high production of reactive oxygen species (ROS), and apoptosis due to increased degradation of pyrophosphate promoting atherosclerotic lesions in vascular wall.^19,30-32^Thus, elevated ALP values in CKD patients may be used as an indicator of declining kidney function as corroborated by Angela Sciacqua et al.^29^ who in their study have shown a significant negative correlation between eGFR and ALP.

However, in stark contrast to the findings of Ray et al.^10^ there is a paradigm shift with respect to serum aminotransferase levels in COVID-19 afflicted CKD patients, which show elevations from baseline levels and lie beyond normal laboratory ranges, this is quite in contrary to that seen in the non-COVID-19 cases with CKD. Mean ALT though elevated, has shown a negative and positive correlation with e-GFR in group 1 and 2 respectively thereby signifying a proportionate fall in enzyme levels with progressive decline in kidney function in Group 2 patients of CKD, with SARS-Cov-2 infection.

## CONCLUSION

Our study is the first of this kind on the Indian population, and most comprehensively describes that SARS-CoV-2 positive CKD patients show more elevations in serum aminotransferase levels as compared to their SARS-CoV-2 positive non-CKD counterparts. Serum ALT values in all SARS-CoV-2 patients show significant correlation (positive or negative) with calculated eGFR values. Thus, ALT may be used as a marker to assess severity of disease and monitor therapeutic response during management of these patients. Also, elevated ALP values in CKD patients may be used as an indicator of declining kidney function. However, more studies in this direction are needed.

## Data Availability

Raw data is available

---

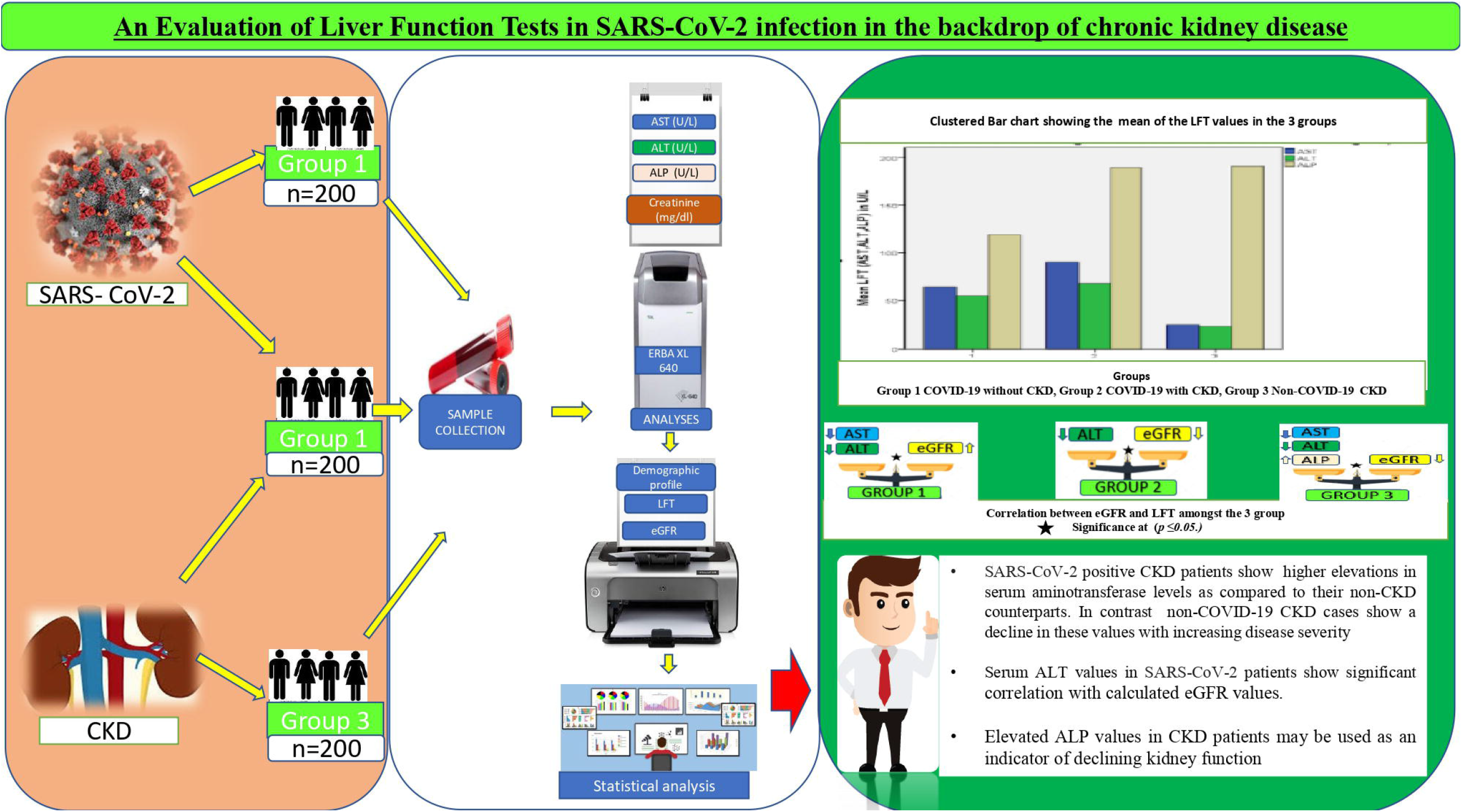

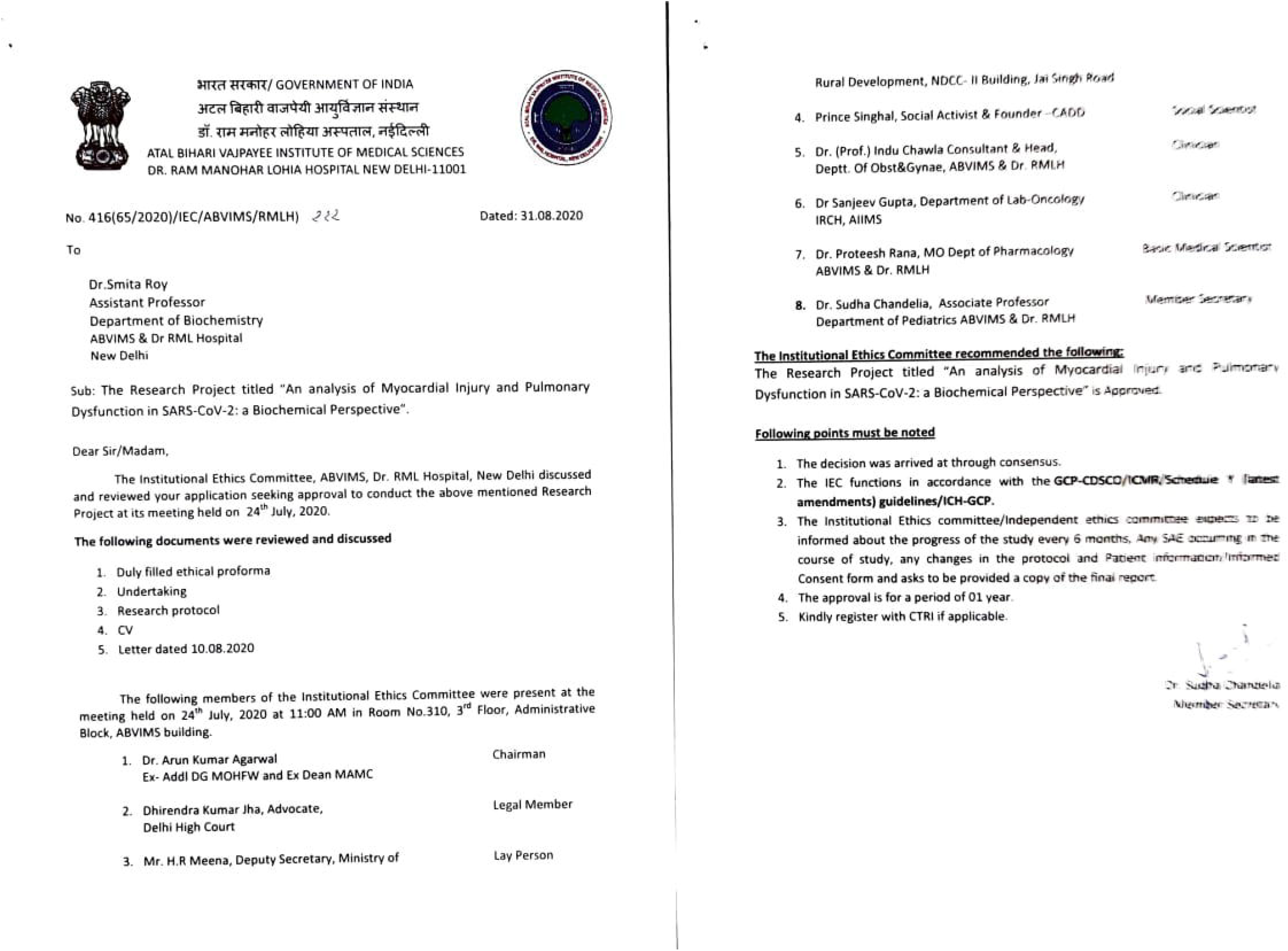

